# Transportation flows and outbreak origins in epidemic spread: Insights from agent-based modeling

**DOI:** 10.1101/2025.10.09.25337674

**Authors:** Konstantin A. Klochkov, Ivan E. Kozlov, Elena N. Ilina, Alexander I. Manolov

**Affiliations:** Research Institute for Systems Biology and Medicine, Moscow, Russia; Moscow Center for Advanced Studies, Moscow, Russia

## Abstract

Human mobility is a key driver of early epidemic spread, and restricting travel remains one of the principal non-pharmaceutical interventions. To better understand how infections propagate through real-world mobility networks, it is essential to disaggregate their components and characterize the functional relationships between mobility flows and epidemic metrics.

Here we introduce transCovasim, an agent-based extension of Covasim that explicitly couples parallel city simulations via inter-city traveler exchange, enabling controlled experiments on mobility and disease dynamics. Using transCovasim, we analyze a two-city system with equal or unequal populations and a hub-and-satellite commuting network parameterized to a Moscow-like setting. In paired identical cities, the mean lag between epidemic peaks scales approximately linearly with the logarithm of inter-city traffic, with steeper delays at lower transmissibility; epidemic variability declines as flows increase. With unequal city sizes, mobility primarily redistributes infections between cities; first-order Sobol’ indices show that peak magnitude is largely insensitive to city’s outbound and inbound flows when sizes are comparable, while sensitivity for a smaller city shifts toward inbound flow as the asymmetry increases. In the hub-and-satellite network, reducing commuting flows before the peak significantly lowers peak incidence, and cumulative infections can still be reduced when restrictions are introduced after the peak; early 100-fold cuts outperform 10-fold cuts, but produce similar results when introduced into the late exponential phase. Finally, dynamic time warping applied to surveillance curves identifies the outbreak’s origin: under Moscow-like flows, accuracy reaches ∼85% by day ∼40 with 10% daily testing, and approaches 100% at lower connectivity. These results clarify how specific mobility patterns shape epidemic timing and burden and provide actionable guidance for mobility-targeted non-pharmaceutical interventions and early source attribution.

**Author summary:** Human mobility governs how epidemics spread between cities, yet policy often treats it as a single lever. We introduce transCovasim, an agent-based extension of Covasim that links parallel city simulations via explicit traveler exchange, allowing controlled tests of pairwise inter-city traffic and hub-and-satellite commuting. In identical cities, the lag between epidemic peaks shrinks approximately linearly with the logarithm of traffic, and stochastic variability decreases as flows rise, especially at lower transmissibility. With unequal sizes, mobility chiefly redistributes infections; sensitivity increases as asymmetry grows. Cutting commuter traffic before the peak reliably reduces peak incidence, while later cuts still lower cumulative burden; early deeper cuts help more. Comparing surveillance curves with time-series alignment can identify the likely source within about a month under moderate testing. These results provide quantitative, network-aware guidance for targeting connections and timing interventions.

## Introduction

Transportation flows play a key role in the early spread of epidemics, and restricting mobility is an important non-pharmaceutical intervention when vaccines or therapeutics are unavailable. High-resolution surveillance during the COVID-19 pandemic provided extensive evidence for the contribution of transport flows to epidemic dissemination at both national and global scales.

A novel coronavirus outbreak (COVID-19) emerged during the Spring Festival travel season (Chunyun), a period when several billion trips occur across China [1]. Stringent restrictions on outbound travel from Wuhan were imposed only about two weeks after the festival began; by then, the estimated probability of at least one imported case exceeded 0.5 in 130 Chinese cities, 107 of which had reported cases by January 26, 2020 [1]. At that point, the risk of importation into Europe, where many countries had yet to detect cases, was already very high [2, 3], although travel curbs reduced international importations from China by roughly 80% by mid-February [4].

COVID-19 initially spread preferentially to cities with large populations and high tourist volumes from Wuhan [5]. During the first wave in mainland China, population mobility from Wuhan was strongly associated with transmission: both cumulative and incident case counts correlated with aggregate traffic flows, especially rail and bus, toward other cities [6–9]. Case numbers in Hubei correlated most strongly with road and rail trips from Wuhan, whereas outside Hubei they correlated more with rail and air travel. Moreover, node centrality metrics in road and air networks (e.g., strength, degree, PageRank) explained cumulative case counts better than direct inflow from Wuhan; for road networks, local centrality (strength, degree) outperformed both flow measures and global centrality (PageRank), consistent with predominantly short-range spread via roads. Epidemic arrival times were most strongly associated with the shortest effective distance from Wuhan [10] in railway networks, somewhat less in road networks, and least in air networks [6]. Taken together, early spread in China was driven primarily by high-volume land transport (rail and bus).

Simulations of the COVID-19 epidemic in England and Wales, launched with different outbreak origins, demonstrate similar qualitative behavior: wave-like spread from urban areas to rural areas [11] (a similar result for measles was obtained by Grenfell et al. [12]). In Europe, five different clusters of temporal dependencies of excess mortality on time during the COVID-19 period are identified, which are associated with specific geographical regions (for example, a difference between western and eastern Europe is noted) [13].

Using global commercial flight data, Stenseth et al. [14] showed that, for the first two months from January 11, 2020, modelled international spread of COVID-19 closely tracked observations until widespread interventions were introduced. In their analysis, reducing transport flows was more effective than entry quarantine, although both were less effective than reducing transmissibility [14]. Complementary modelling for China estimated that non-pharmaceutical interventions, including those targeting transport flows, delayed infection arrival in other cities by a mean of 2.91 days; transport restrictions alone reduced cumulative cases by day 50 nearly fourfold, and by almost 25-fold when combined with other measures [5]. Notably, however, the authors found no clear evidence that restricting traffic outside Hubei reduced case numbers.

Another epidemic in which the role of transportation flows has been systematically examined is influenza.

Pairwise correlations in weekly influenza phases decline with inter-settlement distance and increase with population size; phase correlations also exhibit a nonlinear rise with traffic volume that plateaus at high flow levels [15]. Commuting flows promote synchronization of local epidemics, whereas long-range air travel largely governs international arrival times of pathogens [16]. September international air-traffic volume predicts the timing of the influenza-mortality peak, and the post-9/11 mobility downturn coincided with a delayed season [17, 18]. By contrast, during the 2009 U.S. epidemic, inferred hubs of spread were not the most connected or densely populated areas [19].

Outbreaks seeded in high-density districts (>1,000 persons per km^2^) exhibit substantially less predictable trajectories than those seeded in low-density areas for Ebola, influenza, and measles [20]. In metapopulation models with a generic pathogen and uniform transmissibility and recovery across patches, system stability (eventual fade-out) is governed by average commuting flow rather than by migration per se, although migration can still reshape spatial spread [21]. More broadly, mobility structures transmission, and correlations between inbound and outbound flows can modulate stability conditions [21].

We developed transCovasim, an agent-based epidemiological model built on Covasim [22]. Using this framework, we quantified how the shift in peak infection day between two cities depends on inter-city travel intensity and pathogen transmissibility; assessed sensitivity indices for inbound and outbound flows and insensitivity index.

We further characterized how system metrics change under alternative mobility interventions in hub-and-satellite networks and proposed a method to identify the outbreak’s origin by exploiting temporal offsets between city-level epidemic curves.

## Results

### Two-city transport model with equal agent counts

We considered a two-city model with identical populations (100,000) and symmetric inter-city mobility (equal inbound and outbound flows). A schematic of the setup is shown in Fig 1A. We ran 150 epidemic simulations seeded in one city, varying the daily inter-city traffic (fraction of the population per day) and pathogen infectiousness (fraction of the wild-type SARS-CoV-2 infectiousness).

**Fig 1.**
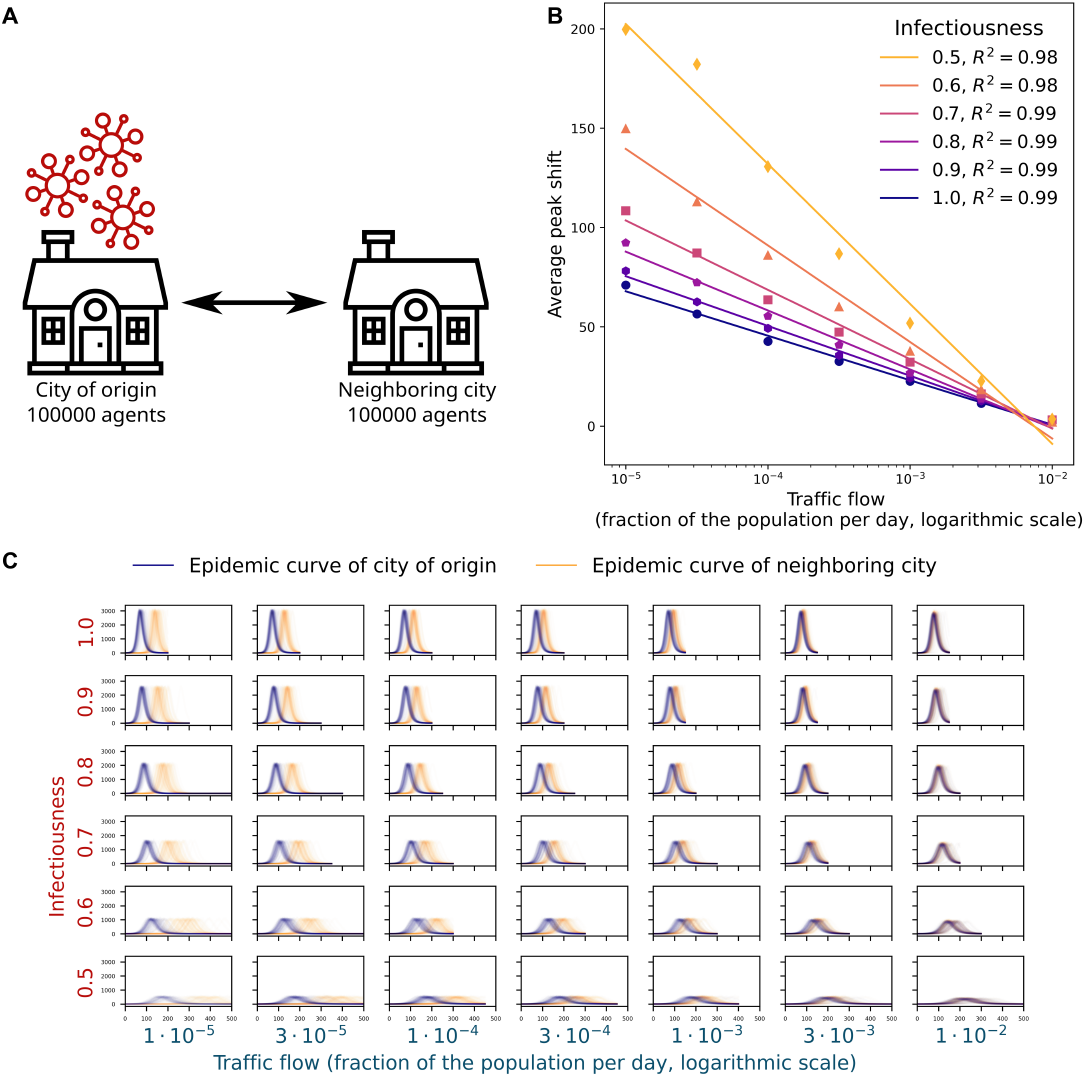
Epidemic spread in a two-city system with identical populations. A: Schematic of the transport model with symmetric inter-city flows. B: Mean offset in the peak-infection day of the destination city relative to the origin city as a function of inter-city traffic (fraction of the population per day; logarithmic scale) across multiple levels of infectiousness (fraction of wild-type SARS-CoV-2 infectiousness). C: Epidemic curves for the origin city (blue) and the destination city (orange) for varying traffic (columns) and infectiousness (rows). Each panel summarizes 150 simulations with outliers removed.

The peak-time offset of the destination city relative to the origin city decreased monotonically with both infectiousness and inter-city traffic, and the result variance declined as well (Fig 1C, S1 Fig).

Fig 1B depicts the mean peak-day offset as a function of traffic flow across infectiousness levels. The offset is well described by a linear relationship with the logarithm of traffic flow, with slope magnitude decreasing as infectiousness increases (coefficient of determination *R*^2^ ∈ [0.98, 0.99]). The t-statistic comparing peak days between the two cities is a non-monotonic, upward-convex function of traffic flow, attaining its maximum at flows below 10^*−*4^. This peak likely reflects discreteness effects when the transported cohort comprises only 1–10 agents. The t-statistic increases systematically with infectiousness (S1 Fig).

### Two-city transport model with unequal populations

We analyzed a two-city model with a fixed total population of 200,000 and varying population ratios (Fig 2A,B). The city of origin had a minimum size of 10,000 agents. In this regime, epidemic metrics depend only on flow from the neighboring city: both cumulative and peak incidence in the origin city increase with inter-city traffic, while they decrease in the neighbor. The peak day in the origin city is essentially invariant to flow, whereas the neighbor’s peak day declines with flow and is downward-convex. Notably, changes in cumulative and peak incidence are of similar magnitude in both cities, indicating that mobility primarily redistributes infections between them (Fig 2A,C).

**Fig 2.**
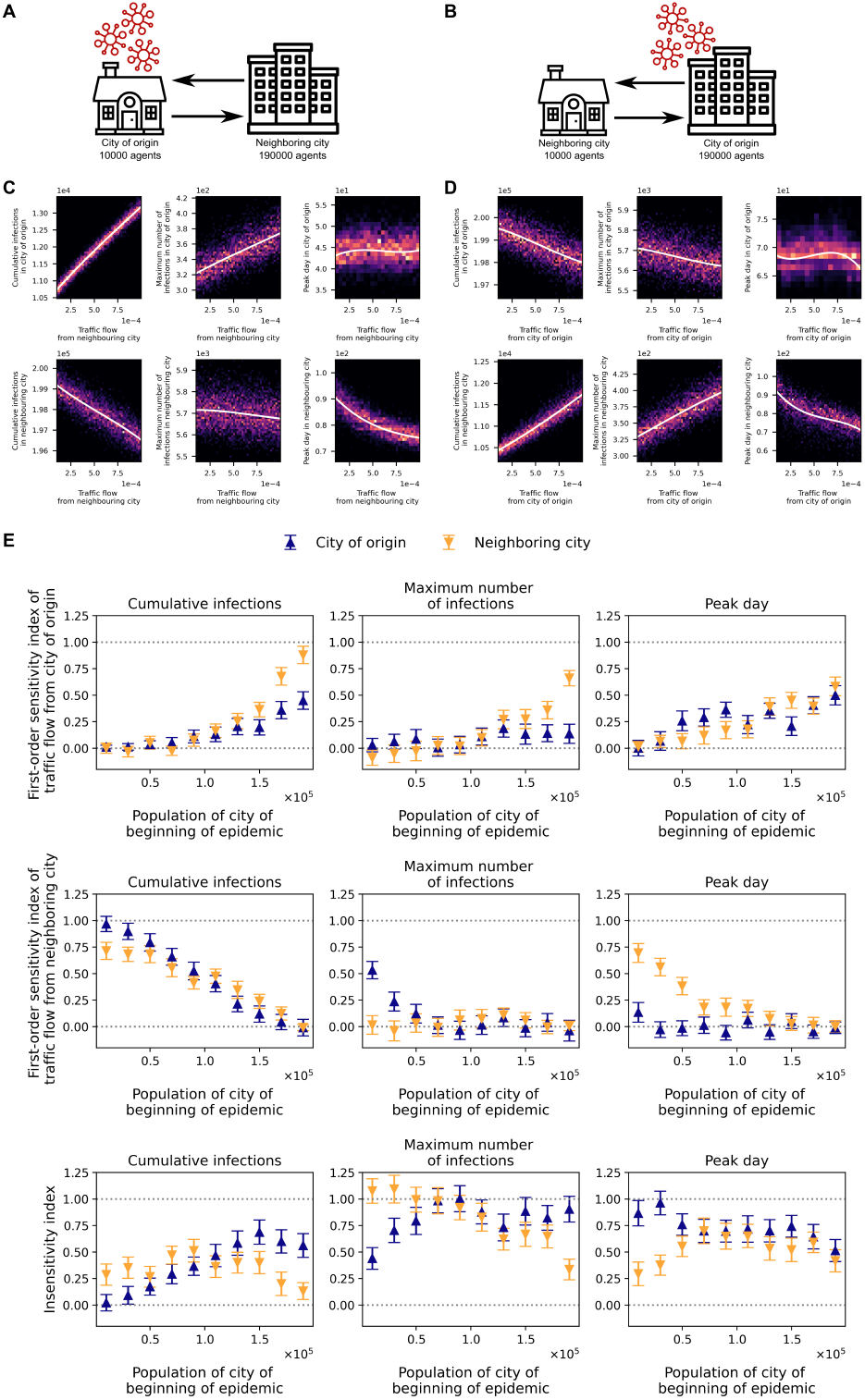
Epidemic spread between two unequal-size cities. A–B: Schematics of the two-city transport model with the outbreak seeded in a city of 10,000 agents (A) or 190,000 agents (B). C–D: Heatmaps of cumulative infections, peak daily infections, and peak-day timing (columns) for the origin city (top rows) and the neighboring city (bottom rows), as functions of (C) flow from the neighbor when the origin has 10,000 agents, and (D) flow from the origin when it has 190,000 agents. E: First-order sensitivity indices versus the city-size ratio for the flow from the origin (top panels), the flow from the neighbor (middle panels), and the resulting insensitivity index (bottom panels), shown for cumulative infections, peak daily infections, and peak-day timing (columns) in the origin city (blue) and the neighbor (orange). Infectiousness is set to the wild-type SARS-CoV-2 level.

At the opposite extreme (origin city size 190,000 agents), the pattern reverses: metrics depend only on flow from the origin, leading to a redistribution of infections toward the neighboring city as traffic increases (Fig 2B,D).

We computed first-order Sobol’ sensitivity indices [23, 24] for the inter-city flow from the outbreak origin and from the neighboring city, across three metrics, cumulative infections, peak incidence, and peak day, in both cities. Indices were estimated using a 4,096-sample Saltelli design [25], with traffic flows varied independently over 10^*−*5^ *−* 10^*−*3^.

Let us denote the first-order Sobol index as S and subtract all first-order Sobol indices from 1, calling this insensitivity and denoting as I (*I* = 1 − ∑ *S*_*i*_). In other words, insensitivity is the proportion of the dispersion of the results that is not associated with the variation of one of the transport flows alone.

As the population of the origin city increases, the first-order sensitivity to outflow from the origin rises, reaching S > 0.8 for cumulative infections in the neighbor and S > 0.4 in the origin, while the sensitivity to outflow from the neighbor declines from S > 0.9 (origin) and S > 0.7 (neighbor) to S < 0.1 for cumulative infections in both cities (Fig 2C,D,E). Thus, cumulative infections in both locations are governed primarily by the absolute number of travelers. For this metric, we observe the lowest insensitivity among those considered: sensitivity increases when city sizes (and hence absolute flows) are highly unequal, and decreases when flows are similar. Sensitivity is highest when the origin is smallest (I < 0.1); when the origin is largest, the neighbor remains sensitive (I < 0.2), whereas the origin becomes comparatively insensitive (Fig 2E).

The maximum number of infections in a city is essentially independent of the outbound tourist flow from that city (S < 0.2) for any population-size ratio. At the same time, the maximum number of infections in the origin city depends only on inbound flow, and only when its population is below 70,000 agents. For the neighbor city, the first-order sensitivity index with respect to the origin’s outbound flow increases as the origin’s population grows (from S < 0.1 to S > 0.6) (Fig 2C,D,E).

This metric is the most insensitive among those considered: when the two cities are similar in size, the insensitivity index exceeds 0.6. A small but noticeable sensitivity appears for the city with the smaller population when the population difference between cities is large (Fig 2E).

The peak infection day in the city of origin is largely insensitive to inflow from the neighboring city (S < 0.2), but it does depend on the origin’s own outflow, although the first-order sensitivity index is non-monotonic (S < 0.5). For the neighboring city, the first-order sensitivity index with respect to flow from the origin increases as the origin’s population grows (from S < 0.1 to S > 0.5), while the index with respect to the neighbor’s own flow decreases (from S > 0.6 to S < 0.1) (Fig 2C,D,E). The insensitivity I of the peak day in the origin city decreases as its population increases, but remains high overall (I > 0.6). In contrast, the neighboring city’s peak-day insensitivity I declines as flows become imbalanced, reaching about I equals 0.3 when the origin city is smallest (Fig 2E).

### Hub-and-satellite transport model

Another common motif in real transport systems is a large city surrounded by smaller, closely connected satellites. We therefore constructed a hub-and-satellite commuting network with one hub and four satellites. To approximate a realistic configuration, the hub-to-satellite population ratio matches that of Moscow and the Moscow region [26], while the total number of agents is set to one-twentieth of the real population to reduce computation time. Inter-satellite flows were generated using a gravity model [27–29], with traffic proportional to city populations and inversely proportional to the square of inter-city distance. A schematic of the resulting transport model is shown in Fig 3A.

**Fig 3.**
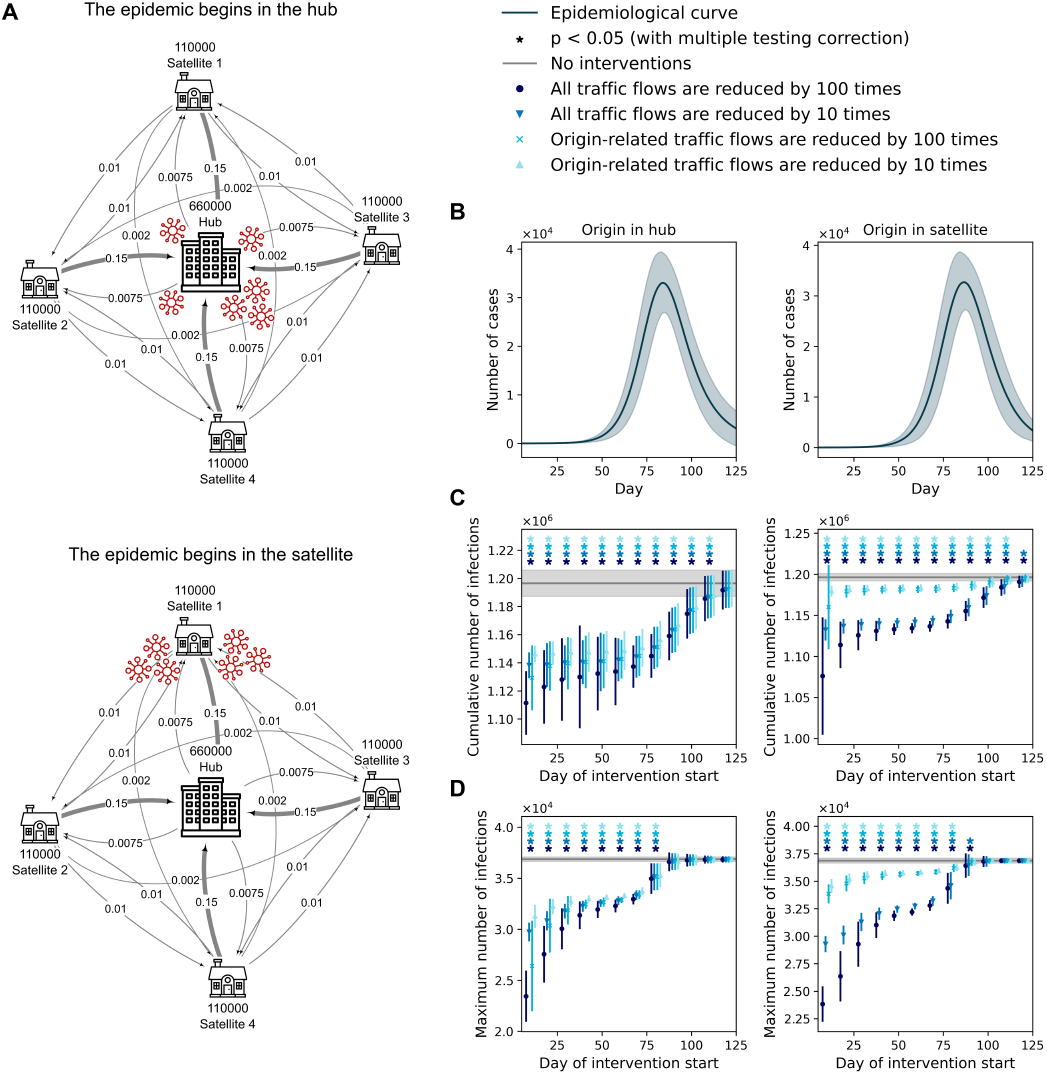
Hub-and-satellite transport model. A: Schematic of the hub-satellite system represented as a graph, with outbreaks initiated in the hub (top) or in a satellite (bottom). Vertex labels indicate city population sizes (number of agents), and edge labels indicate the daily commuting fraction of the population. B: Epidemic curves for outbreaks starting in the hub (left) or in a satellite (right). Shaded areas denote the standard deviation across 150 simulations. C–D: Cumulative infections (C) and peak infections (D) in the hub-satellite system as functions of the day interventions in commuting flows are introduced, for outbreaks starting in the hub (left) or in a satellite (right). Curves of different colors correspond to different transport-flow interventions. The gray curve indicates the mean across 150 baseline simulations without interventions; shaded areas show standard deviations. Stars mark statistically significant differences (Student’s t-test, p < 0.05 with multiple-testing correction). Infectiousness is set to the wild-type SARS-CoV-2 level.

We ran 150 simulations with the outbreak seeded in either a satellite or the hub (Fig 3A). We then introduced mobility interventions at various times: a 10-fold or 100-fold reduction of all flows in the network or only the flows directly connected to the origin city. For comparison, an additional 150 baseline simulations were run without restrictions. Outcomes were summarized by two metrics: cumulative infections and peak daily infections.

The left panels of Fig 3C,D show how the system metrics vary with the timing and type of intervention when the outbreak starts in the hub. The gray line indicates the mean (with standard deviation) from 150 simulations without restrictions. An asterisk marks time points where the metric under restrictions differs significantly from the no-restriction baseline (Student’s t-test, significance level 0.05 with multiple-comparisons adjustment). For context on intervention timing, Fig 3B plots the epidemic curves when the outbreak originates in the hub (left) or in a satellite (right).

When the outbreak begins in the hub, imposing restrictions up to the peak day yields a statistically significant reduction in peak infections (Fig 3D). Cumulative infections decline significantly even when restrictions are introduced after the peak and maintained through day 120, by which time daily incidence has fallen to about 13% of its maximum (Fig 3B,C). During days 40–70 (late exponential phase), the effects of 10-fold and 100-fold flow reductions are similar, whether applied network-wide or only on hub-satellite links, and the effect size remains essentially stable across the end of the exponential phase (Fig 3C,D). Even with interventions introduced at this stage, cumulative infections decrease by 3.8% (*p <* 10^*−*98^) and peak infections by 8.5% (*p <* 10^*−*184^) (Table 2). By contrast, the gap between 10-fold and 100-fold restrictions is most pronounced early in the epidemic: a 100-fold reduction lowers peak values by an additional factor of roughly 1.5-2 compared with interventions initiated at the end of the exponential phase (Table 1).

**Table 1.** The magnitude of the reduction in target metrics when introducing transport restrictions on the 10th day of the epidemic.

|  | Origin in hub |  |  |  | Origin in satellite |  |  |  |
| --- | --- | --- | --- | --- | --- | --- | --- | --- |
|  | Restriction on all flows |  | Restriction on hub-related flows |  | Restriction on all flows |  | Restriction on satellite-related flows |  |
|  | 100 times | 10 times | 100 times | 10 times | 100 times | 10 times | 100 times | 10 times |
| Cumulative infections reduction | 7.1% | 4.9% | 5.6% | 4.2% | 10.1% | 5.4% | 3.0% | 1.4% |
| Maximum infections reduction | 36.4% | 19.3% | 28.4% | 15.2% | 35.4% | 20.6% | 8.2% | 6.2% |

**Table 2.** The magnitude of the reduction in target metrics when introducing transport restrictions on the 70th day of the epidemic.

|  | Origin in hub |  |  |  | Origin in satellite |  |  |  |
| --- | --- | --- | --- | --- | --- | --- | --- | --- |
|  | Restriction on all flows |  | Restriction on hub-related flows |  | Restriction on all flows |  | Restriction on satellite-related flows |  |
|  | 100 times | 10 times | 100 times | 10 times | 100 times | 10 times | 100 times | 10 times |
| Cumulative infections reduction | 5.0% | 4.4% | 4.3% | 3.8% | 5.0% | 4.4% | 1.2% | 1.1% |
| Maximum infections reduction | 10.6% | 9.5% | 9.4% | 8.5% | 11.0% | 9.9% | 2.7% | 2.4% |

We observe analogous patterns when the outbreak starts in a satellite (right panels of Fig 3C,D; Tables 1, 2). The key difference is that restricting only the flows incident to that satellite is far less effective than network-wide restrictions, although both produce statistically significant reductions in the target metrics.

### Detecting the outbreak’s origin in the hub-satellite model

Experiments were performed on the hub-satellite model described above, varying infectiousness and a global multiplier applied to all traffic flows. For each combination, 150 simulations were run; results are summarized in Fig 4. When the outbreak starts in the hub, satellite curves are markedly more synchronous than when the outbreak starts in a satellite, an effect evident even at a flow multiplier of 0.3. Subtle desynchronizations are more clearly revealed in phase portraits (Fig 4B). These observations motivate detecting the outbreak’s origin via temporal dependencies between city-level trajectories.

**Fig 4.**
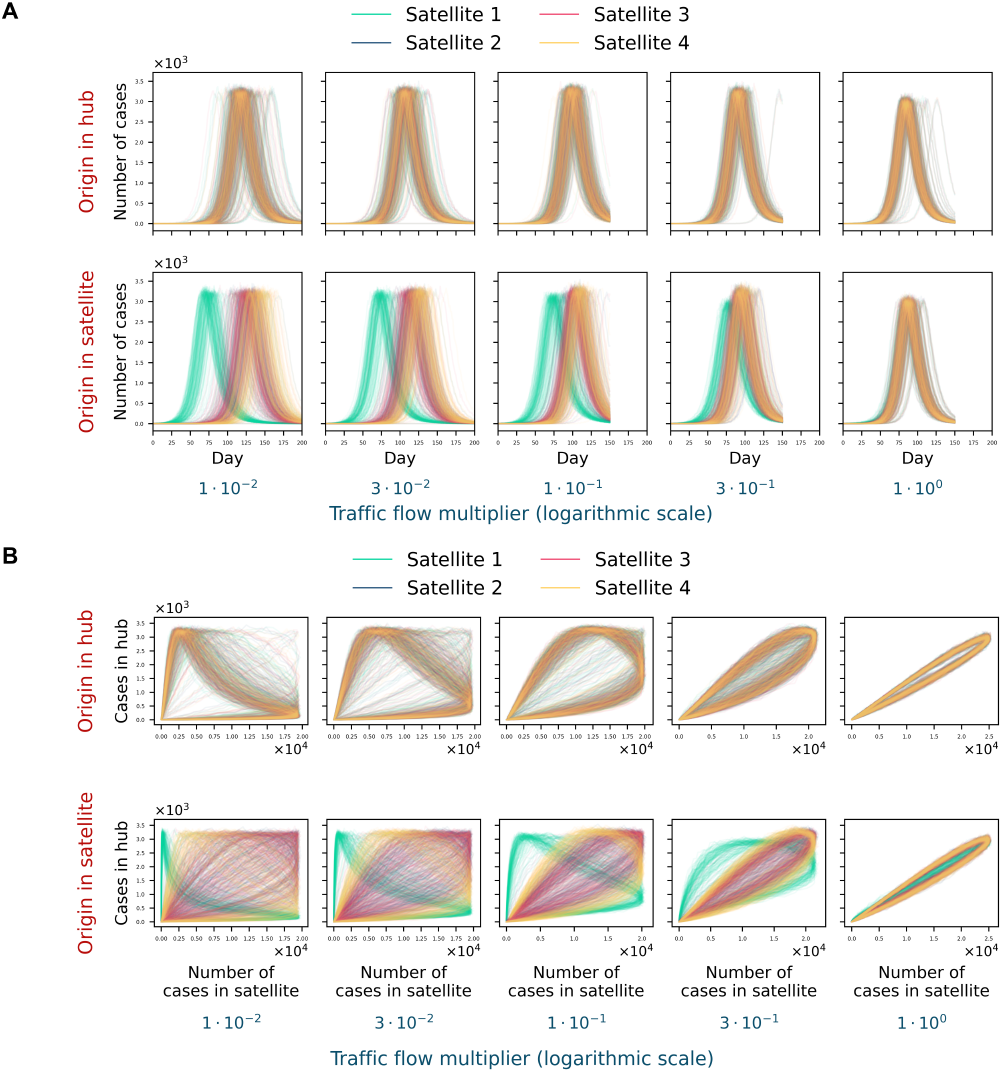
Relative shifts among satellite epidemic curves in the hub-satellite system. A–B: Satellite incidence curves (A) and phase portraits (B) for outbreaks seeded in the hub (top rows) or in a satellite (bottom rows), shown across traffic-flow multipliers (panel columns) with infectiousness set to the wild-type SARS-CoV-2 level. Phase portraits highlight temporal offsets that are less apparent in raw incidence curves.

To infer the outbreak’s origin from temporal dependencies, we applied dynamic time warping (DTW) to the hub’s incidence curve and each satellite’s curve [30]. From the DTW alignment, we computed the mean temporal offset between each pair of curves by averaging the pointwise time shifts along the optimal path. The curve with the earliest inferred onset (smallest mean shift) was designated as the origin city; in the event of ties, one of the tied cities was selected at random (Fig 5A). To approximate real-world conditions, we incorporated agent testing (see Methods).

**Fig 5.**
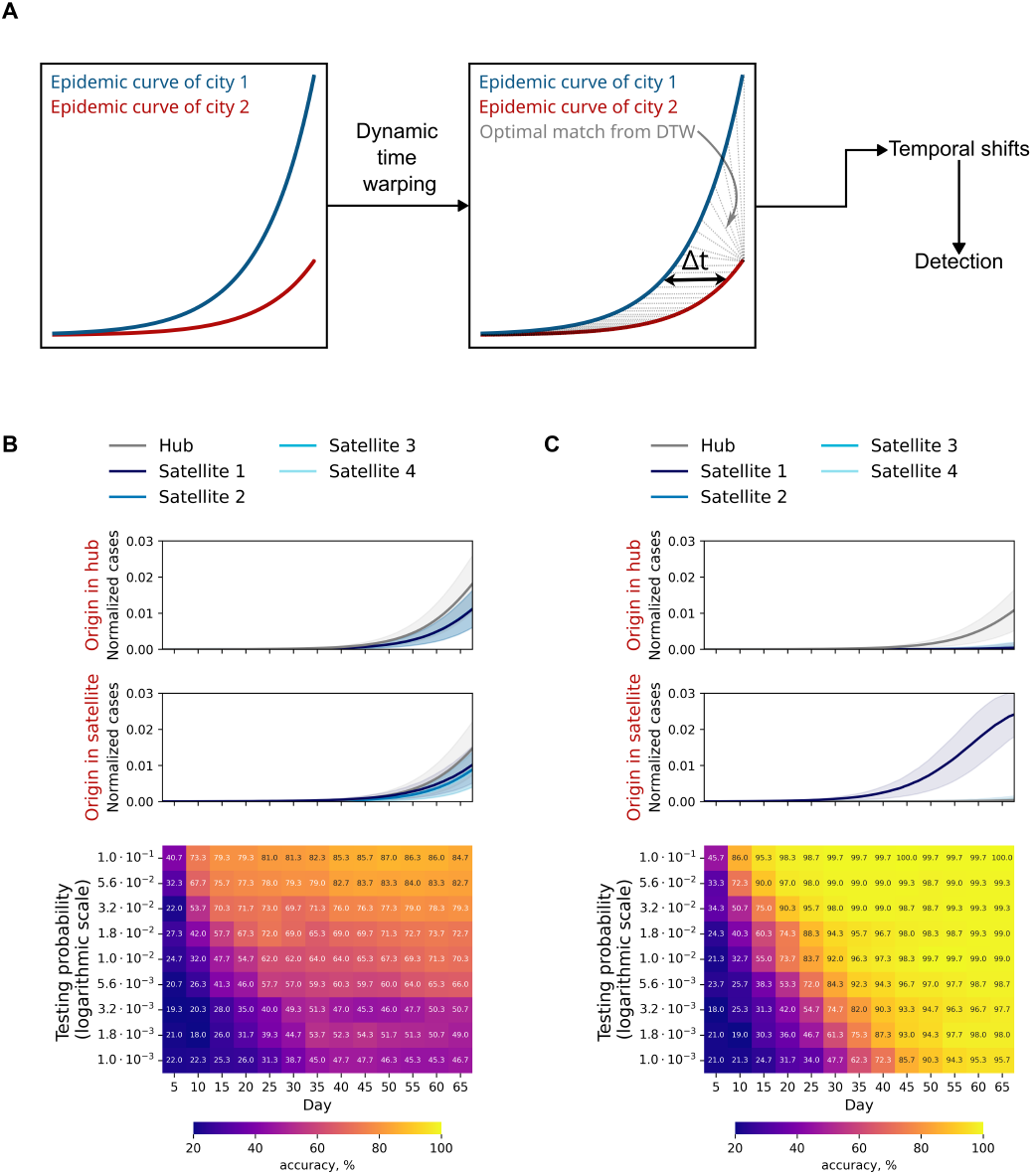
Detecting the outbreak’s origin in the hub-satellite model. A: Schematic of the DTW-based method for identifying the origin city. B-C: Heatmaps of origin-detection accuracy versus epidemic day for systems with traffic flows equal to (B) or 100-fold smaller than (C) those of Moscow and the Moscow region. Testing is simulated by sampling detected infections from a binomial distribution.

With Moscow-region-like flows, testing more than 2% of the population yields about 70% origin-detection accuracy by day 25, after which accuracy plateaus due to high synchrony. With lower testing fractions, the maximum attainable accuracy falls to roughly 50%. The best performance in this scenario is about 85% when testing 10% of the population, reached by day 40 at the latest (Fig 5B).

For hub-satellite networks with weaker connectivity, the maximum detection accuracy approaches 100% across all tested sampling fractions. However, in the time-testing plane, the heatmap reveals a region where reliable detection is impossible because too few infections are identified at those epidemic times (Fig 5C, S2 Fig).

At lower infectiousness, the boundary of the heatmap’s nondetection region shifts to later epidemic times; at infectiousness equal to 0.5 of the wild-type SARS-CoV-2 level, this boundary is delayed by roughly twofold.

## Conclusion and discussion

In this study, we analyzed two foundational transport-network configurations, the two-city system and the hub-satellite system, that underlie more complex mobility structures.

For two identical cities, we found a linear relationship between the neighbor’s peak-time shift (relative to the outbreak city) and the logarithm of inter-city traffic, with the slope magnitude decreasing as transmissibility increases. This supports the use of logarithmic effective distances [6, 10] when a single characteristic travel mode dominates, in our experiments, movements with mean duration 7 days, 40 contacts, and a transmissibility multiplier equal to that of random contacts in Covasim. In practice, this suggests such distances are appropriate on temporal or spatial scales where one mode prevails (e.g., air travel on an international scale, or rail during China’s Spring Festival).

We also observed a upward-convex relationship between the t-statistic comparing peak days in the two cities and the inter-city traffic flow. The presence of an extremum suggests a discreteness effect at flows corresponding to roughly 1–10 travelers per 100,000 residents per day, where result variance increases markedly.

Notably, the t-statistic’s dependence on transmissibility is the inverse of the peak-shift trend: as transmissibility rises, the mean peak shift shrinks while its variability shrinks even more, yielding a larger t-statistic.

In the two-city system with unequal sizes, cumulative infections in each city become more sensitive to traffic flows as size asymmetry and passenger volumes increase, and they depend on the absolute number of travelers. By contrast, when cities are similar in size, the peak infections are largely independent of flows; with large size disparities, the smaller city’s peak becomes flow-sensitive. The neighbor’s peak day depends on the absolute number of travelers, whereas the origin’s peak day is sensitive only to changes in its own outbound flow. Overall sensitivities are modest: for many city-size ratios, the first-order sensitivity to any single flow does not exceed 0.25, implying that no more than one quarter of the variance in a given metric is attributable to variation in that flow alone.

In the hub-satellite system with an outbreak seeded in the hub, 10-fold and 100-fold reductions applied either network-wide or only on hub-linked flows yield statistically significant decreases in peak and cumulative infections when introduced by the peak and maintained through day 120, respectively. When implemented at the end of the exponential phase, all four restriction types perform similarly. By contrast, early in the epidemic, a 100-fold reduction achieves roughly 1.5-2 times greater peak reduction than a 10-fold reduction. Analogous patterns hold when the outbreak starts in a satellite, but restricting only satellite-related flows is notably less effective than network-wide measures. These results underscore the value of timely non-pharmaceutical interventions in hub-based networks and the need to target the hub itself regardless of the outbreak’s point of origin.

We evaluated source-city detection feasibility in the hub-satellite system as a function of transport-flow magnitude and daily testing fraction. For Moscow-region-like flows, the maximum achievable accuracy is about 85% when testing 10% of the population per day; reducing flows drives accuracy toward 100%. Accuracy heatmaps (testing fraction vs. epidemic day) reveal a parameter region where detection is impossible because too few infections are identified during that period. As transmissibility decreases, the boundary of this nondetection region shifts to later epidemic days, by roughly twofold at transmissibility equal to half the wild-type SARS-CoV-2 level. At the same time, increased relative variability in case counts at lower transmissibility degrades detection quality. This helps explain why, under Moscow-like flows, accuracy typically does not exceed about 80%, except at the 10% testing level where it can peak near 85%. Achieving 80% accuracy across testing levels appears feasible when flows are below roughly 3% of the real values at transmissibility equal to 0.5 of the wild-type SARS-CoV-2.

However, our study has several limitations. Analyses on real transport networks should distinguish among road, bus, rail, and air flows, since network structure and epidemiological impact differ markedly across modes [6, 8, 16]. Trip purpose (e.g., commuting vs. tourism) determines duration and thus transmission potential [20], as does the volume of random movements [31]. In practice, tourists often travel in family groups rather than as independent individuals, and travel exhibits spatial-temporal regularity [32] — features not yet represented in transCovasim. Incorporating such detail, especially at country scale, would require extensive, personalized mobility data (e.g., mobile-operator records or survey data) [33] and remains an important direction for future work.

## Methods

### Agent-based model

transCovasim is built on Covasim [22], an open-source agent-based model that incorporates realistic population structure, disease progression, and interventions.

The transCovasim model makes the following assumptions:

1. Agents move instantaneously between cities;
2. Agents in severe, critical, or dead states do not travel;
3. No infections occur during the instantaneous movement step;
4. Travelers are sampled uniformly at random from the origin population;
5. Upon arrival, travelers form random contacts within the destination city.

During initialization, each city is assigned a traveler-capacity equal to twice the expected number of travelers in that city, so, by default, the capacity is not saturated. Potential traveler agents are preallocated to each city as “absent” so that array sizes remain fixed during simulation, preserving performance, and they are configured to participate in social interactions per the specified parameters.

After initialization, the model proceeds through the main simulation loop. At each time step, a random set of traveler agents is drawn in each city according to the prescribed flows. These travelers are temporarily removed from their home city’s contact network and inserted into the destination city’s network, where they engage in a fixed number of random contacts. After the trip duration elapses, the process is reversed. Agents in severe, critical, or dead states do not travel: if selected to travel, they remain in their home city; if they enter such a state while away, they return to the home city only after recovery. The core algorithms in Covasim’s simulation loop are implemented as highly optimized operations on 32-bit Numba arrays [34]. For efficiency, agents are not instantiated as individual objects but as slices across state arrays.

To enable transport flows, we augmented Covasim’s state-array schema with four agent-level attributes:

1. `trueId`: original identifier in the home city, used to restore identity upon return;
2. `inCity`: boolean flag indicating whether the agent is currently present in the city;
3. `restInAnotherCityDays`: number of days remaining in the current stay outside the home city;
4. `ownCity`: identifier of the agent’s home city.

In the current model, the following traffic-flow parameters are configurable:

1. `adjacencyMatrix`: city-city flow matrix; each entry is the daily fraction of the origin city’s population traveling to the destination;
2. `timeRelax`: mean trip duration in days (default: 7);
3. `interventionData`: transport-intervention spec; a dictionary indicating which links are restricted, the start day, duration, and the reduction factor applied to flows;
4. `contactCount`: mean number of contacts per traveler per day with residents of the destination city (default: 40);
5. `beta`: multiplier for transmission probability on the traveler layer (default: 0.3).

A detailed description of the algorithms is provided in the supplementary material (S1 File).

### Design of the experiments

Computational experiments were conducted in Python. For each parameter set, we ran 150 simulation replicates with different random seeds. Metrics are reported as the mean across 150 runs, with standard deviations indicating variability.

In the two-city experiments, the mean stay in the destination city was 7 days, the number of contacts was 40, and the transmissibility multiplier for travelers was 0.3 (as for random contacts in [22]). For the identical-cities scenario, we excluded outliers before further analysis, specifically, simulations in which the epidemic failed to start or did not spread to the neighboring city.

Sobol’ sequences and first-order sensitivity indices were computed using SALib [35, 36].

In the hub-satellite experiments calibrated to Moscow and the Moscow region, the region was partitioned into four satellite cities. Bidirectional flows between the hub and satellites were set from mobile-operator data [26]. For simplicity, we used a square geometry with the hub at the center: the distance between neighboring satellites is 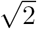 times the hub-satellite distance, and between opposite satellites is 2 times the hub-satellite distance. Trip duration was 1 day, the number of contacts was 40, and the traveler-layer transmission multiplier was 0.6, consistent with short commuter trips to workplaces in neighboring cities.

In the detection experiments, testing was simulated by sampling detected cases from a binomial distribution whose success probability equaled the true proportion of new infections.

## Data availability statement

All data and codes created and analyzed in this study are available in the GitHub repository at the following link: https://github.com/KonstantinKlochkovv/transportation-motifs

## Supporting information

**S1 Fig.**
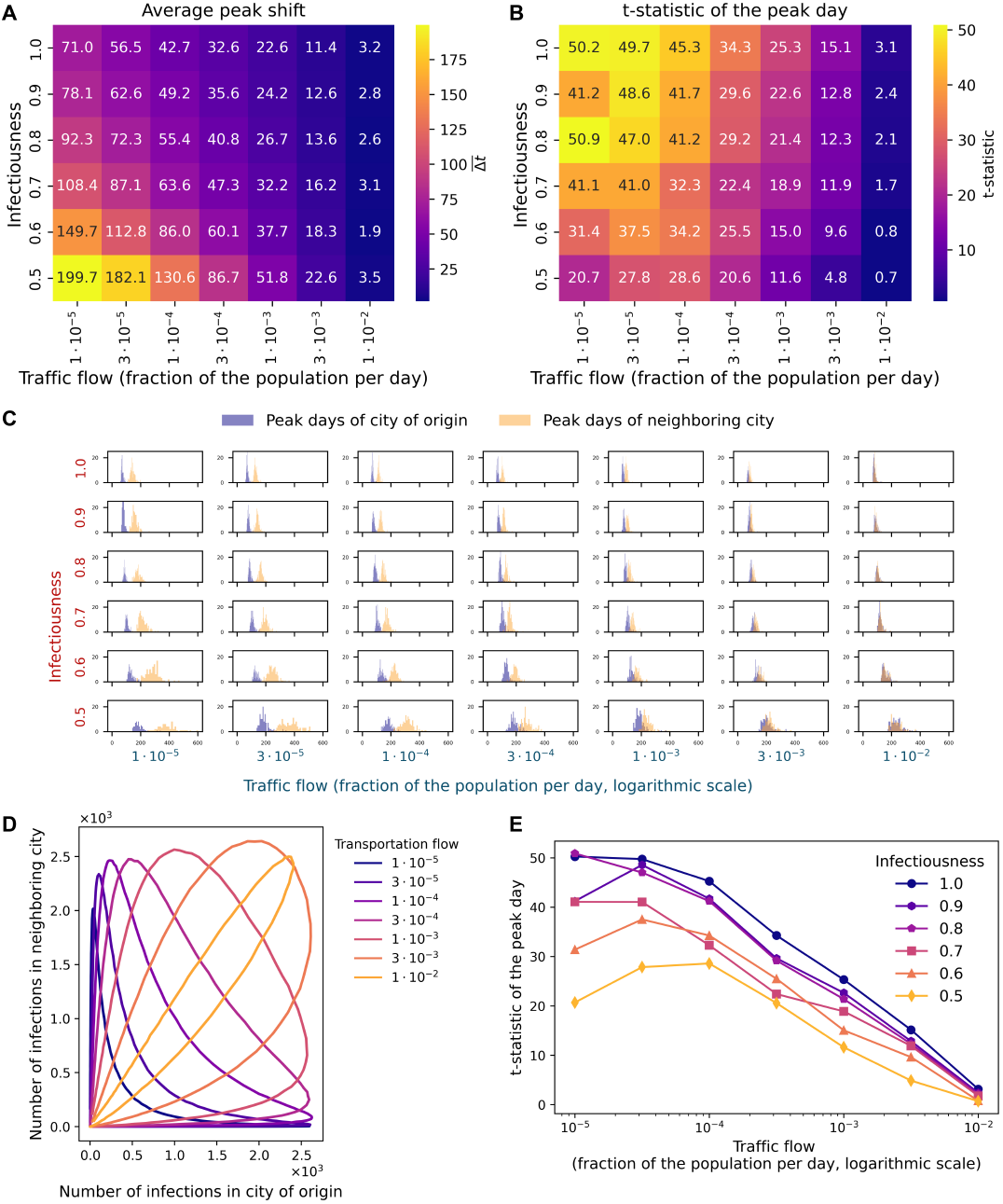
Epidemic spread in a two-city system with identical populations. A–B: Heatmaps of the mean offset in the peak-infection day of the destination city relative to the origin city (A) and of the t-statistic comparing peak days (B), based on the transport flow between cities (fraction of the population per day) at different infectiousnesses (fraction of the infectiousness of the wild variant SARS-CoV-2). C: Histograms of the distribution of the peak infection day in the city of origin (blue) and in a neighboring city (orange) at different levels of traffic flow between cities (columns) and infectiousnesses (rows). D: Phase portrait of the two-city system at different traffic flows between them at the infectiousness of the corresponding wild variant SARS-CoV-2. E: Dependence of the t-statistic when comparing the days of peak infections in cities in the system on the traffic flow between cities at different infectiousnesses. Each panel shows the result of 150 simulations after removing outliers.

**S2 Fig.**
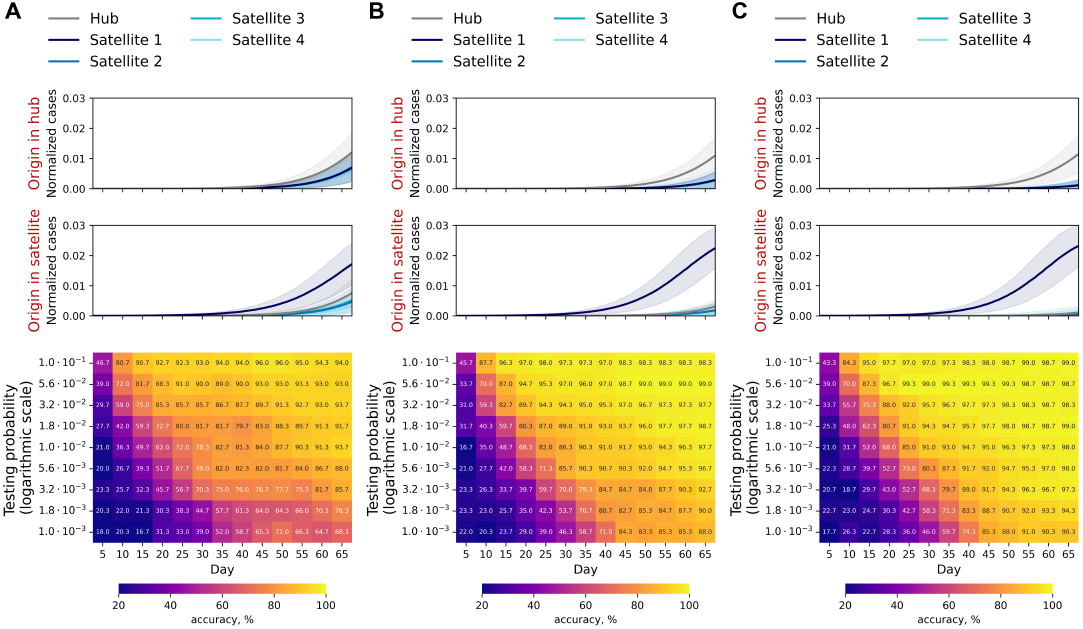
Detecting the outbreak’s origin in the hub-satellite model. A–C: Heatmaps of origin-detection accuracy versus epidemic day for systems with traffic flows 3.16 (A), 10 (B), and 31.6 (C) times smaller than those of Moscow and the Moscow region. Testing is simulated by sampling detected infections from a binomial distribution. Infectiousness set to the wild-type SARS-CoV-2 level.

**S1 File. Descriptions of the algorithms underlying the transCovasim model**.

### Initialization

During the initialization phase, the number of places allocated for tourists is determined for each city as a fraction of the city’s population equal to

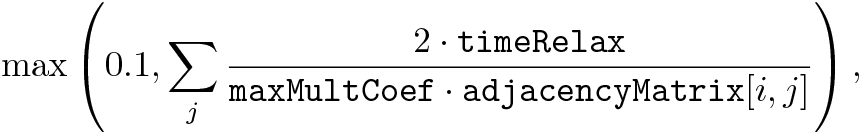

In other words, the number of places allocated to tourists is greater than or equal to twice the mathematical expectation of their number, with the default value of `maxMultCoef` equal to one.

These agents are then added to cities as absent (`inCity = False`) and integrated into social interactions using `touristLayer` in accordance with the specified parameters.

After initialization, the model enters the main simulation cycle.

### Function for placing people in the destination city

The function passes all attributes of tourist agents `rest` to agents in the destination city with indices `inds`, except for the identifier `uid`, and then marks their presence in the city (*people*.*inCity*[*inds*] *← True*) (Algorithm 1).

### Function for placing people who should return to the city

The function returns agents `backPeople` according to their native identifier `backPeople.trueUid` using the function `UpdatePeopleByRest`.

#### Algorithm 1

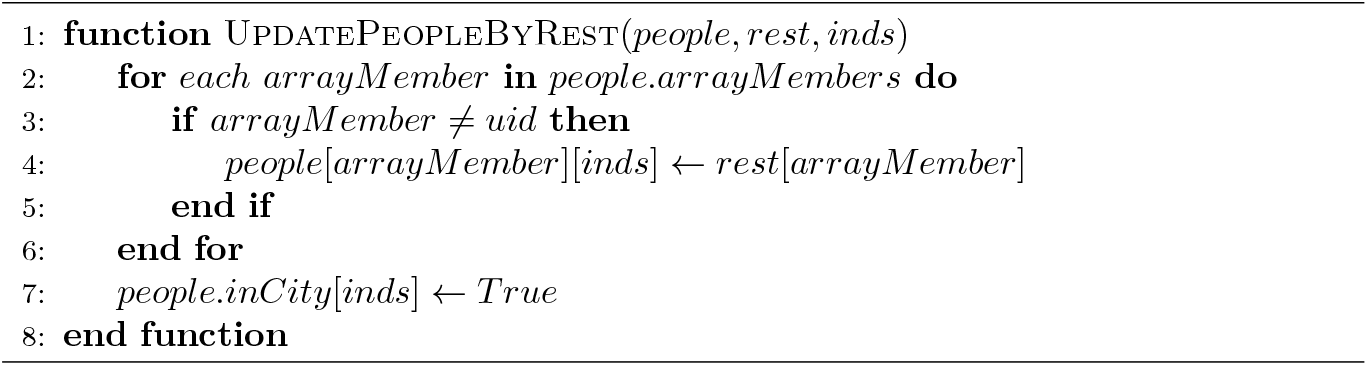

### Function for placing tourists who arrived in the city

The function determines the identifiers available for tourist accommodation in the destination city, selects the required number of them corresponding to the number of arriving tourists, if there are enough places, or all remaining places otherwise. Then, agents are placed in the places with the selected identifiers using the `UpdatePeopleByRest` function (Algorithm 2).

#### Algorithm 2

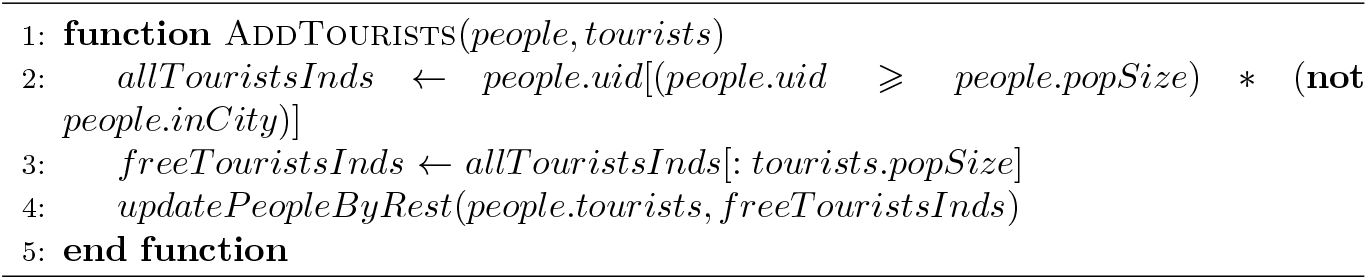

### Function for choosing and extracting tourists who should leave their city

The function corresponding to the daily outflow of tourists from the city `outflowRatioToCities` sets their numbers from a Poisson distribution. It is checked whether there are enough people in the city to send. If there are not enough, the number of tourists to be sent is recalculated according to the number of people in the city. Then, this number of agent identifiers is selected from among the local residents (`people.uid` < `people.popSize`) who are in the city (`people.inCity == True`). This list is divided into lists by destination city based on known transport flow ratios `outflowRatioToCitiesPercent`. For each destination city cityInd, severe, critical, and dead agents are removed from the list of tourists. The remaining agents are assigned a duration of stay in another city `people.restInAnotherCityDays`, which is either identically equal to 1 for all when modeling commuting flows, or from a Poisson distribution without 0 otherwise. Then, all departing agents from the `touristPeople` list are extracted from the departure city and stored as a list of pairs (`cityInd, touristPeople`) (Algorithm 3).

### Function for choosing and extracting people who should return to their city

The function determines the indices of agents whose home city matches the index in the cycle (`people.ownCity == cityInd`), whose travel time has expired (`people.restInAnotherCityDays == 0`), and who are not severe, critical, or dead at the current simulation step. After that, agents with these indices `backPeople` are extracted from the city where they are located and added to the list of returns as a pair (`cityInd, backPeople`) (Algorithm 4).

#### Algorithm 3

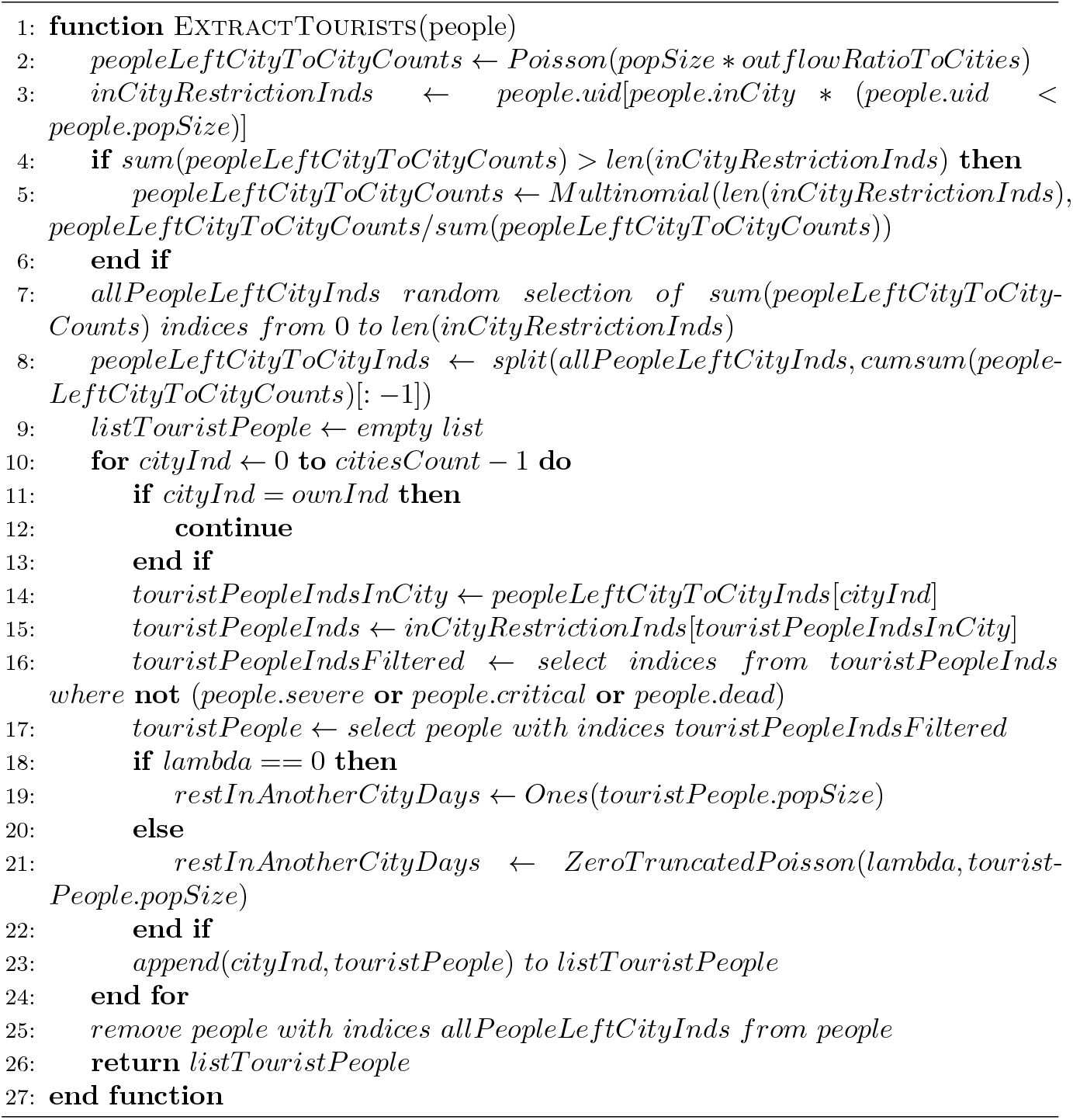

#### Algorithm 4

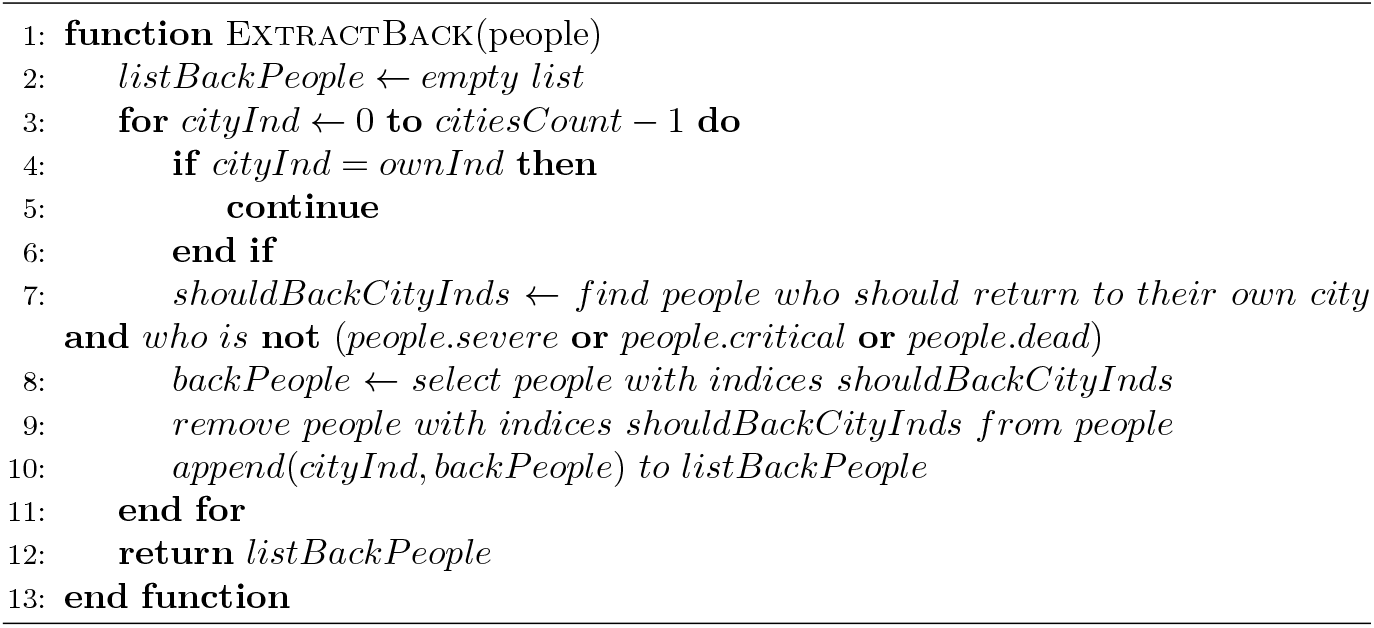

